# Towards accurate and unbiased imaging based differentiation of Parkinson’s Disease, Progressive Supranuclear Palsy and Corticobasal Syndrome

**DOI:** 10.1101/19007575

**Authors:** Marta M Correia, Tim Rittman, Christopher L Barnes, Ian T Coyle-Gilchrist, Boyd Ghosh, Laura E Hughes, James B Rowe

## Abstract

The early and accurate differential diagnosis of parkinsonian disorders is still a significant challenge for clinicians. In recent years, a number of studies have used MRI data combined with machine learning and statistical classifiers to successfully differentiate between different forms of Parkinsonism. However, several questions and methodological issues remain, to minimise bias and artefact-driven classification. In this study we compared different approaches for feature selection, as well as different MRI modalities, with well matched patient groups and tightly controlling for data quality issues related to patient motion.

Our sample was drawn from a cohort of 69 healthy controls, and patients with idiopathic Parkinson’s disease (n=35, PD), Progressive Supranuclear Palsy Richardson’s syndrome (n=52, PSP) and corticobasal syndrome (n=36, CBS). Participants underwent standardised T1-weighted MPRAGE and diffusion-weighted MRI. We compared two different methods for feature selection and dimensionality reduction: whole-brain principal components analysis, and an anatomical region-of-interest based approach. In both cases, support vector machines were used to construct a statistical model for pairwise classification of healthy controls and patients. The accuracy of each model was estimated using a leave-two-out cross-validation approach, as well as an independent validation using a different set of subjects.

Our cross-validation results suggest that using principal components analysis (PCA) for feature extraction provides higher classification accuracies when compared to a region-of-interest based approach. However, the differences between the two feature extraction methods were significantly reduced when an independent sample was used for validation, suggesting that the principal components analysis approach may be more vulnerable to overfitting with cross-validation. Both T1-weighted and diffusion MRI data could be used to successfully differentiate between subject groups, with neither modality outperforming the other across all pairwise comparisons in the cross-validation analysis. However, features obtained from diffusion MRI data resulted in significantly higher classification accuracies when an independent validation cohort was used.

Overall, our results support the use of statistical classification approaches for differential diagnosis of parkinsonian disorders. However, classification accuracy can be affected by group size, age, sex and movement artifacts. With appropriate controls and out-of-sample cross validation, diagnostic biomarker evaluation including MRI based classifiers can be an important adjunct to clinical evaluation.

## Introduction

The early and accurate differentiation of parkinsonian disorders poses a challenge for clinicians and trialists, which will become critical with the advent of disease modifying therapies (van Eimeren et al., 2019). Early symptoms of akinetic rigidity and non-motor symptoms often overlap between idiopathic Parkinson’s disease (PD), progressive supranuclear palsy (Richardson syndrome, PSP) and degenerative corticobasal syndrome (CBS, and its pathological counterpart corticobasal degeneration, CBD). PD is the most common form of parkinsonism, with approximately 140 cases per 100,000 (Porter et al., 2006) whereas PSP and CBS are each approximately 3 per 100,000 (Coyle-Gilchrist et al., 2016). Misdiagnosis of PSP and CBS is common, often as PD, taking on average nearly three years from initial symptoms to diagnosis, while many cases remain undiagnosed.

There is a pressing need for reliable biomarkers to differentiate these disorders, not only to aid diagnosis in early or atypical cases, but to monitor progression in trials and to support *ante mortem* studies of pathogenesis (van Eimeren et al., 2019). Biomarkers should be objective and observer-independent, reproducible, informative about the underlying biology and ideally non-invasive. Candidate biomarkers for parkinsonian disorders have included cognitive tests (Aarsland, 2003; Pillon et al., 1995; Rittman et al., 2013) and assays of cerebral spinal fluid, serum or urine such as neurofilament-light (Jabbari et al., 2017; Constantinescu et al., 2019), supplementing those clinical features that have high clinicopathological correlations (Alexander et al., 2014; Gazzina et al., 2019; Respondek et al., 2017).

Magnetic Resonance Imaging (MRI) provides a set of potential biomarkers (Whitwell et al., 2017), with the advantages of being non-invasive, widely available and versatile. Multiple MRI methods have the potential to inform about the underlying neural systems and the changes resulting from specific pathologies. Pathognomic radiological signs have been reported, such as the “mickey mouse” and “hummingbird” signs of midbrain atrophy in PSP, but such abnormalities are insensitive, especially in early stage disease when there would be most to gain from disease modifying therapies. Moreover, visual assessment of MRI images is dependent on the experience of the observer.

Automated methods have been developed using volumetric or intensity change in grey matter (GM), for example voxel based morphometry (VBM). Most VBM studies of grey matter in degenerative parkinsonian syndromes have compared patients to healthy controls (Beyer, Janvin, Larsen, & Aarsland, 2007; Brenneis et al., 2004; Cordato, Duggins, Halliday, Morris, & Pantelis, 2005; Ghosh et al., 2012; Summerfield et al., 2005; Yarnall et al., 2014), whereas few have compared different patient groups against each other (Boxer et al., 2006; Price et al., 2004). Other studies have compared subgroups within each disorder, according to cognitive impairment (Mak et al., 2015; Paviour, Price, Jahanshahi, Lees, & Fox, 2006) or neuropsychiatric symptoms (Ghosh et al., 2012; Yao et al., 2014). White matter (WM) changes have also been described, using VBM or diffusion tensor imaging (DTI) measures such as the fractional anisotropy (FA) and mean diffusivity (MD). Differences are observed for PD patients vs controls (Goveas et al., 2015; Rae et al., 2012; Yoshikawa, Nakata, Yamada, & Nakagawa, 2004; K. Zhang et al., 2011), PD vs PSP (Seppi et al., 2003) and PD vs CBS (Boelmans et al., 2010). A meta-analysis of 43 DTI studies in parkinsonian syndromes (Cochrane & Ebmeier, 2013) suggested the potential of diffusion-weighted imaging to improve the differential diagnosis of parkinsonism. However, accuracy was often not greater than clinical criteria, sample sizes were often small, and the utility for single subject decision-making was limited.

Here we propose that better classification can be achieved by alternative approaches to magnetic resonance imaging data, using statistical classifiers such as support vector machines (SVM). Mulitvariate data features from a training set of data (subjects) can be used to build a model to classify a new dataset (one or more new subjects). In addition to individual subject classification, these methods can identify which features underlie the classification (i.e. indicative of relevant pathological features) and indices of confidence or typicality that could be used to assess progression. Statistical classifiers have been successfully applied to a number of neurological and psychiatric disorders, including schizophrenia (Caan et al., 2006; Ingalhalikar, Kanterakis, Gur, Roberts, & Verma, 2010), Alzheimer’s disease, frontotemporal dementia (Davatzikos, Resnick, Wu, Parmpi, & Clark, 2008) and autism spectrum disorder (Bloy et al., 2011; Ingalhalikar et al., 2010; Ingalhalikar, Parker, Bloy, Roberts, & Verma, 2011). Haller and colleagues (Haller et al., 2012) used DTI data from 17 PD patients and 23 patients with other forms of “atypical parkinsonism” (including typical PSP and multisystem atrophy). Using tract based spatial statistics (TBSS), a non-linear SVM algorithm, and a 10-fold cross-validation, classification between PD and other patients was accurate (97.5 ± 7.5%, depending on the number of features used for model training). In combination with manual regions-of-interest selection, classification accuracies >95% were also achieved by Proedoehl and colleagues (Prodoehl et al., 2013) in binary differentiation of PD and PSP. T1-weighted MRI can also support binary classification, >85% (Focke et al., 2011; Salvatore et al., 2014). Unfortunately, whilst previous studies have demonstrated success for differential diagnosis in parkinsonism, significant limitations and methodological questions remain. First, many studies have used poorly matched groups in terms of age or clinical variables, and most studies have used different numbers of subjects in each group. The latter is of particular concern because commonly used statistical classifiers which minimize the classification error, such as support vector machines, are liable to inflate accuracy from unbalanced datasets (see for example (He & Garcia, 2009; Tang, Zhang, & Chawla, 2009)).

A second problem relates to the selection of features used by the classifier. For example, previous studies have used either mean values from specified regions or individual voxel data, including manual selection with its operator dependence. In addition, studies have rarely compared MRI modalities to assess whether T1-weighted or diffusion-weighted images (DWI) are most useful for differential diagnosis of movement disorders.

A third problem concerns the validation of results, which is challenging with small group sizes. Most studies have included small numbers of subjects, and therefore employed cross-validation techniques. However, the use of the same subjects for training and validation is controversial and may inflate classification accuracies. A more conservative approach is to split the data in two independently acquired groups: one for training and the other for validation (Salvatore et al., 2014).

Finally, most studies have failed to consider how different levels of motion during the MRI acquisition affect classification accuracies. This issue is particularly important when working with patients with movement disorders. Head motion results in artefacts and smoothing of MRI data. Different levels of motion across groups could significantly contribute to classifier’s apparent success in separating patient groups.

In the present study we aimed to address these four methodological issues in the context of differential diagnosis of PD, PSP and CBS. Specifically, we compare three equal-sized and closely-matched groups of patients; we used automatic feature selection of grey and white matter signals; and we undertook an initial leave-two-out cross-validation followed by validation in an independent data set. The comparison of well-matched groups, with automatic feature selection is a challenge for imaging markers, but one that is necessary to develop unbiased and useful clinical research tools.

## Methods

### Subjects

Our analysis sample was drawn from a cohort of 69 healthy controls (mean age 67.3 years, range 51 to 84), 35 people with idiopathic PD (mean age 66.9 years, range 46 to 76, UK Parkinson’s disease brain bank criteria), 52 people with probable PSP (mean age 71.9 years, range 51 to 92, MDS clinical diagnostic criteria for PSP-Richardson’s syndrome (Höglinger et al., 2017)), and 36 people with probable CBS (mean age 66.9 years, range 39 to 88, (Armstrong et al., 2013)). A neurologist experienced in movement disorders undertook the UPDRS-III motor subscale for all patients. For the cross-validation analysis (see below), 19 cases per group were selected so as to match for age, sex, MRI motion, and similar UPDRS-III score in the patient groups. Local Ethical Committee approval and written informed consent were obtained. All participants had mental capacity to consent under UK law.

### MRI data acquisition

Diffusion and T1-weighted MRI data were acquired for all subjects using a 3T Siemens Tim TRIO scanner at the Wolfson Brain Imaging Centre. Diffusion MRI data was acquired with a twice refocused spin echo (TRSE) sequence (Reese, Heid, Weisskoff, & Wedeen, 2003). Diffusion sensitising gradients were applied along 63 non-collinear directions with a b-value of 1000s/mm^2^, together with one acquisition without diffusion weighting (b=0). The remaining imaging parameters were: TR=7800 ms, TE=90ms, matrix=96×96, field of view (FoV)=192×192 mm, slice thickness=2 mm without gap, interleaved slice acquisition, and the PAT mode was GRAPPA with an acceleration factor of 2. A high resolution 3D T1-weighted MPRAGE image was also acquired (TR=2300 ms, TE=2.98 ms, FOV=256×240 mm, matrix=256×256, slice thickness=1 mm).

### Quality assurance and exclusion criteria

MRI data in general, and diffusion MRI in particular, can suffer from significant distortions in the presence of head motion. Given the motor deficits associated with parkinsonism, metrics of motion are especially important to ensure the quality of the data across control and patient groups. Estimating the amount of motion in 3D MPRAGE images is not trivial. We used SPM12 (www.fil.ion.ucl.ac.uk/spm/) to estimate the level of smoothness associated with the MPRAGE images of each subject. While not a direct measure of head motion artefacts, the inherent smoothness in the data correlates with motion. Firstly we performed full image segmentation using the *Segment* tool in SPM12 (Ashburner & Friston, 2005). Secondly, the *spm_estimate_smoothness* function was used to estimate the inherent smoothness associated with soft tissue outside the brain, cerebral spinal fluid (CSF) and bone. This function returns a spatial smoothness estimator based on the variances of the normalised spatial derivatives as described in (Kiebel, Poline, Friston, & Holmes, 1999). The estimated smoothness values were then compared across controls and patients, and significant outliers (>2 standard deviations from the mean) were removed from further analysis.

For the diffusion MRI data we estimated motion artefacts in two ways. Firstly, we used the *eddy_correct* function in FSL v5.0.9 (www.fmrib.ox.ac.uk/fsl) to perform affine registration between each diffusion weighted volume and the b=0 image. The output log files from *eddy_correct* were used to estimate the absolute displacement between each diffusion MRI volume and the b=0 images, as well as the relative displacement between a given volume and its predecessor. Significant outliers (>2 standard deviations from the mean) on either metric were identified and removed from further analysis. Subjects were also excluded if they moved more than 3mm (1.5 × voxel size) between any two diffusion MRI volumes. Secondly, we used an automated method for detection of striping patterns in the data (Neto-Henriques, Cam-CAN, & Correia, 2016). Stripping artefacts are caused by spin history and are a common consequence of head motion when interleaved MRI acquisitions are used. Subjects with more than five volumes affected by stripping artefacts were excluded.

### Cross-validation and validation groups

The remaining subjects were divided into two subgroups: a cross-validation group and an independent validation group. The subjects included in the cross-validation group were selected to satisfy the following criteria:

- Equal numbers of subjects across the four control/patient groups
- No significant differences in motion metrics across the four control/patient groups
- No significant age or sex differences across the four control/patient groups
- UPDRS-III scores matched for all three patient groups

All remaining subjects who had not been excluded by the motion quality control metrics made up the validation group.

### Pre-processing of MRI data

The T1-weighted MPRAGE images were segmented and normalised into MNI space using SPM12. Firstly, the MPRAGE images were segmented into grey and white matter maps using *Segment* (Ashburner & Friston, 2005). For this step, six tissues types were considered (grey matter, white matter, CSF, bone, soft tissue outside the brain, and air and other signals outside the head). Segmentation was then followed by DARTEL (Diffeomorphic Anatomical Registration Through Exponentiated Lie Algebra) (Ashburner, 2007), an algorithm which increases the accuracy of inter-subjects alignment by modelling the shape of each brain using three parameters per voxel, and generating an increasingly sharp average template over several iterations. Finally, the sixth iteration of the DARTEL template was used to generate spatially normalised and Jacobian scaled grey matter images in MNI space (Ashburner, 2009; Mechelli, Friston, Frackowiak, & Price, 2005).

The diffusion MRI data were skull-stripped and motion corrected using FSL v5.0.9, and the diffusion tensor model fitted using a non-linear fitting algorithm implemented in C and matlab. Fractional Anisotropy (FA) and mean diffusivity (MD) were computed for each subject. FA and MD maps were transformed onto a common template space using DTI-TK, a tensor-based registration approach (Hui Zhang et al., 2007; H Zhang, Yushkevich, Alexander, & Gee, 2006), and a study-specific population based atlas (Hui Zhang, Yushkevich, Rueckert, & Gee, 2007).

### Feature extraction

For the GM maps obtained from segmentation of T1-weighted images, feature extraction was performed in two ways: (A) using the cortical and subcortical regions-of-interest from the Harvard-Oxford Atlas (http://neuro.imm.dtu.dk/wiki/Harvard-Oxford_Atlas), and (B) using principal component analysis (PCA).

For the region-of-interest analysis, 63 grey matter cortical and subcortical ROIs were applied to the spatially normalised GM maps for each subject, and the average GM density value per ROI calculated, hence generating 63 independent features per subject (Figure 1A). For the PCA analysis, a GM mask was first created by thresholding the GM template obtained from DARTEL. This mask was then applied to the images from each subject, and the voxels contained within the mask were included in a multi-subject PCA analysis, resulting in *N-1* independent features, where *N* represents the number of subjects included in this analysis (Figure 1B).

**Figure 1.**
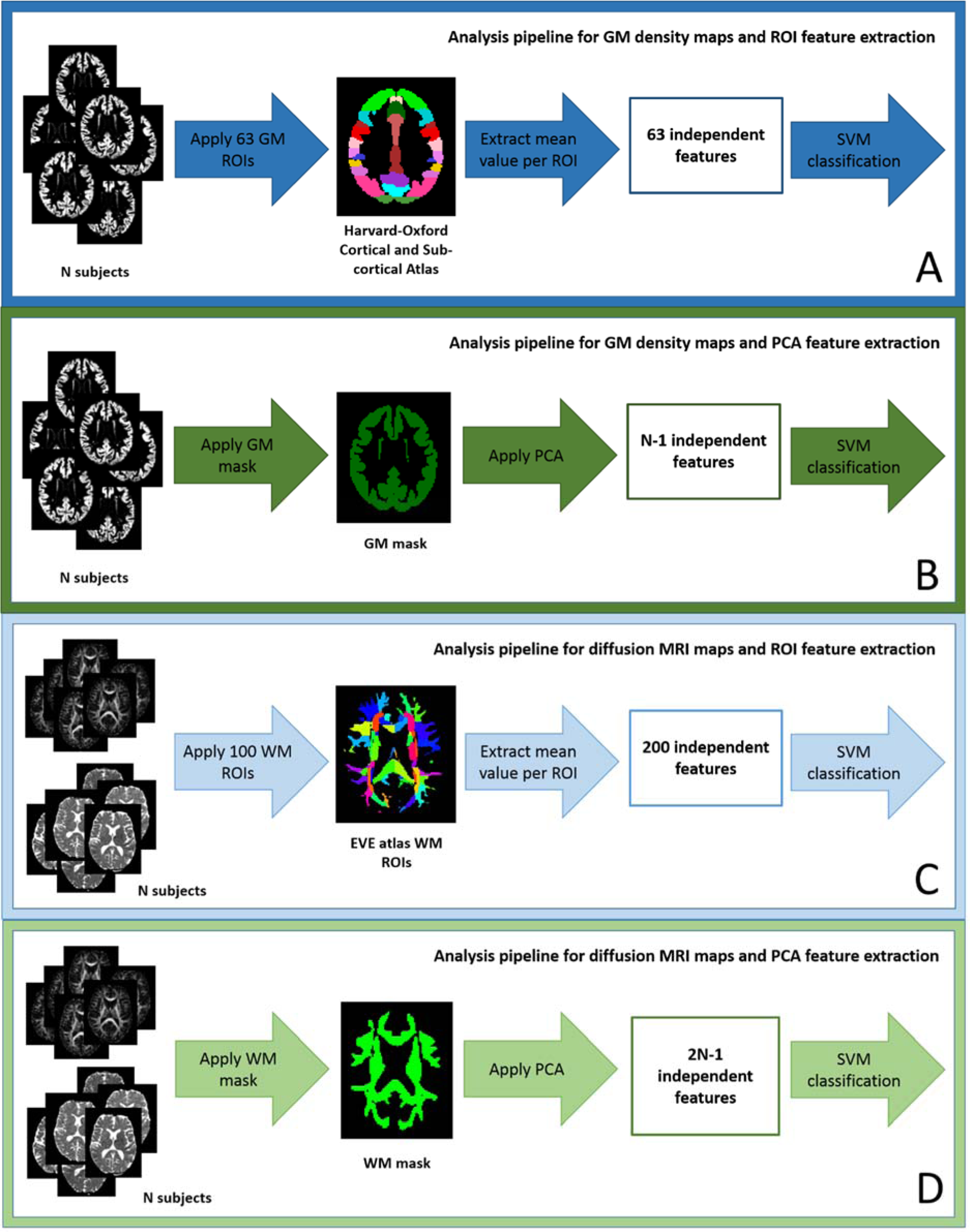
Analysis pipeline for each combination of data type and feature extraction method. (A) T1-weighted MRI and ROIs. (B) T1-weighted MRI and PCA. (C) Diffusion MRI and ROIs. (D) Diffusion MRI and PCA.

The same two methods for feature extraction were applied to the FA and MD maps. For the ROI approach, the white matter regions from the EVE atlas (http://lbam.med.jhmi.edu/) were used to extract the average FA and MD values for each region and subject (Figure 1C). For PCA, a white matter mask was first generated by thresholding the FA map corresponding to the study-specific template. Voxels selected from the FA and MD maps of all subjects were included to generate *2N-1* independent features (Figure 1D).

### Feature ranking and statistical classification

Four parallel streams of subsequent analysis were performed, one for each data type and feature extraction method combination: (A) GM maps + ROIs, (B) GM maps + PCA, (C) diffusion maps + ROIs, and (D) diffusion maps + PCA.

Following feature extraction, the features generated by each approach were ranked separately, using the Fisher Discriminant Ratio (FDR):

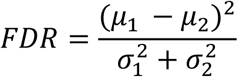

where *μ*_*i*_ and 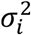 denote the mean and the variance of the *i*-th class, respectively.

The top feature for each stream was used in combination with support vector machines (SVMs) to construct a statistical model for pairwise classification of healthy controls, PD, CBS and PSP. The remaining features were added to the model, one at a time, in the order of their FDR ranking, and the classification accuracy of each model as a function of the number of features was calculated. The SVM analysis was performed using the LIBSVM package in matlab (Chang & Lin, 2011).

To assess the accuracy of the four analysis streams, we first used leave-*n*-out cross-validation, in view of the modest sample size of the well-matched groups. The *N* available subjects are randomly split into a training set of size *(N-n)* and a test set of size *n*. In this study, n=2 for the pairwise comparisons, with the testing set including one subject from each group. The training set is used to build a model and the validation of the resulting classifier is performed on the training set. Multiple rounds of cross-validation are then performed for different permutations of the two subjects left out of the training set. The average classification accuracy across all iterations of the cross-validation process is reported. However, this method may inflate classification accuracies. Therefore the leave-two-out cross-validation was supplemented by an independent validation using a different set of cases altogether when estimating the model’s accuracy. For the cross-validation approach, feature ranking using FDR was recalculated for each fold using the subjects in the training subgroup only, and the same ranking applied to the two subjects left out. For the independent validation, the FDR ranking was determined using the cross-validation group only, and the ranking order applied to the subjects in the validation group.

### Data availability

Participant consent prevents open data access but academic (non-commercial) requests for data sharing would be welcome. Please contact the senior author. The principal softwares used (SPM, FSL, libsvm and Matlab) are publically available.

## Results

### Quality assurance and subject exclusion

Figure 2 shows examples of MRI images for the subjects excluded by the motion quality control assessment. Our exclusion criteria reduced the sample size to 62 controls (7 subjects excluded by DWI motion metrics), 32 PD (1 subject excluded by DWI motion metrics, 2 subjects excluded by both DWI and MPRAGE metrics), 33 PSP-Richardson’s syndrome (16 subjects excluded by DWI motion metrics, 3 subjects excluded by both DWI and MPRAGE metrics) and 26 CBS (6 subjects excluded by DWI motion metrics, 4 subjects excluded by both DWI and MPRAGE metrics). Overall, the PSP group was the most affected by motion, with a total of 19 subjects excluded.

**Figure 2.**
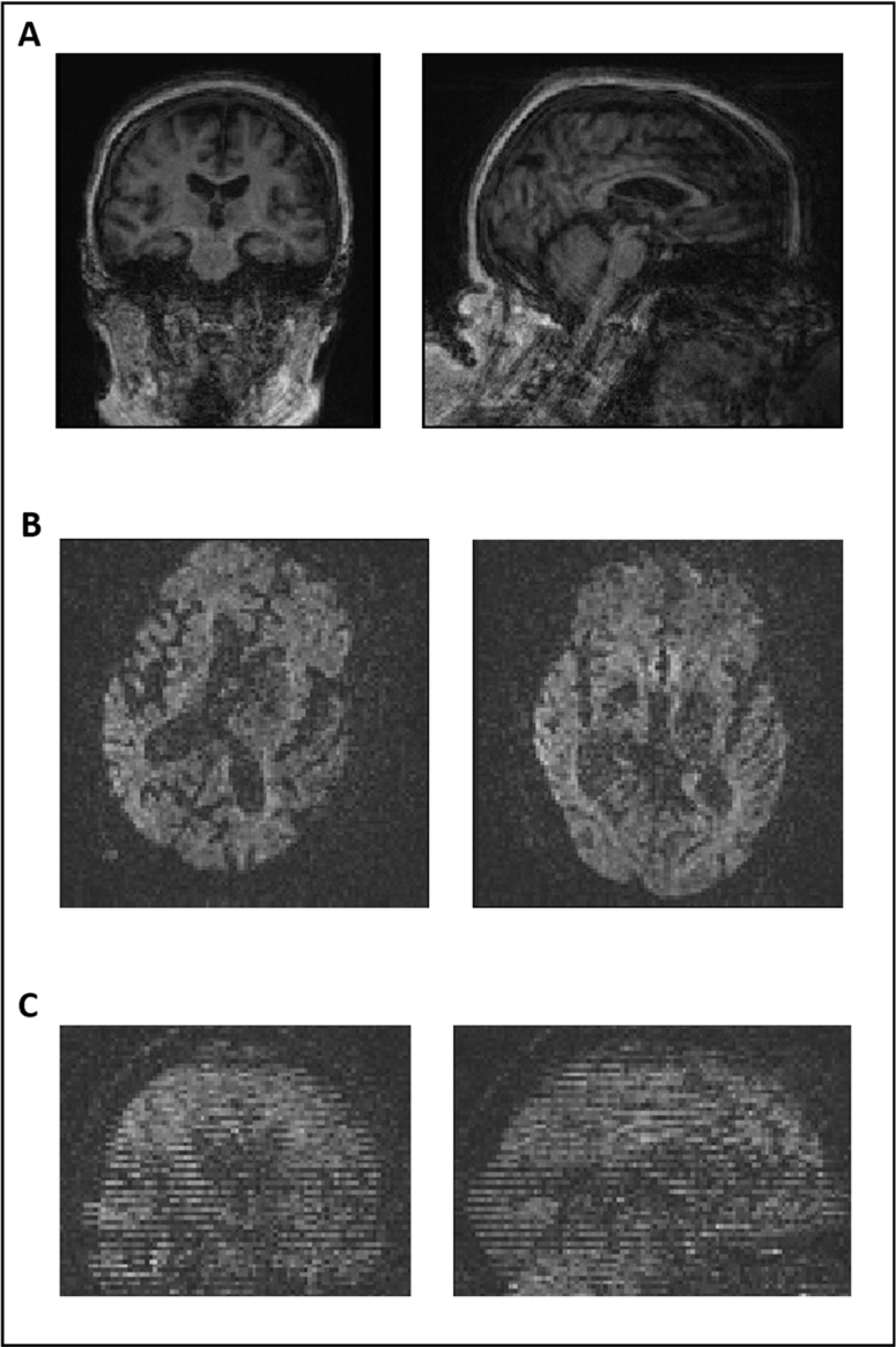
Examples of MRI images for subjects excluded by the motion quality control procedures. (A) T1-weighted MPRAGE for a PSP patient identified as outlier by the estimated smoothness of the segmented soft tissue outside the brain. (B) Two consecutive slices for the diffusion MRI data for a PSP patient identified as an outlier by the absolute and relative displacement metrics. (C) Diffusion MRI data for a CBS patient identified by the automated stripe detection algorithm.

### Cross-validation and validation groups

After quality assurance, patient groups were confirmed to be matched for motion metrics, age and sex using ANOVA or chi-squared, as appropriate. However, the 62 controls were younger than the patients, and included a larger proportion of females. To remove these confounding differences, we randomly removed females and younger subjects to reach a sample of 43 age- and sex-matched healthy controls.

To form the cross-validation group, 19 patients were selected with each diagnosis, matching demographics and UPDRS-III, with 19 controls matching them for motion metrics, age and sex. The remaining 58 subjects (24 controls, 13 PD, 14 PSP and 7 CBS) formed the independent validation test cohort.

Table 1 shows the demographic and neuropsychological evaluation scores for all groups. Motion quality control metrics are also shown. For the cross-validation group, there was no significant difference by diagnosis in sex, age or UPDRS-III score. There was a significant difference in MMSE score across the different groups, and post-hoc tests revealed that both PSP and CBS patients had a lower MMSE score when compared to healthy controls and PD patients. All motion metrics were matched across groups. For the independent validation group age and head motion were matched across groups, but there were mild differences between PSP and controls or CBS in terms of age or smoothness respectively (see Table 1).

**Table 1.** – Demographic, neurophysiological evaluation scores and quality control information. Data are shown as mean ± standard error (range). ^a^Chi-squared test, ^b^ANOVA, ^c^ANOVA, ^d^ANOVA followed by post hoc tests (Control>CBS p<0.01, Control>PSP p<0.05, PD>CBS p<0.01 and PD>PSP p<0.05), ^e^ANOVA, ^f^ANOVA, ^g^ANOVA, ^h^Chi-squared test, ^i^due to the different samples sizes pairwise comparisons were conducted using t-tests (PSP>CBS p=0.035, all other comparison p>0.05), ^j^due to the different sample, sizes pairwise comparisons were performed using t-tests (Control<PSP p=0.011, all other comparison p>0.05), ^k,l^due to the different sample sizes pairwise comparisons were conducted using t-tests, all of which resulted in p>0.05.

|  | Controls | PD | PSP | CBS | Difference between groups |
| --- | --- | --- | --- | --- | --- |
| Cross-validation group |  |  |  |  |  |
| sample size | 19 | 19 | 19 | 19 | -- |
| sex f/m (%) | 36.8/63.2 | 47.4/52.6 | 42.1/57.9 | 52.6/47.4 | $p=0.786^a$ |
| age (years) | 66.2±1.6<br>(54.9-81.1) | 65.0±1.9<br>(46.9-76.9) | 69.1±1.3<br>(60.8-83.2) | 68.2±2.3<br>(39.1-88.2) | $p=0.373^b$ |
| UPDRS-III score | -- | 20.5±2.1<br>(5-32) | 27.2±3.4<br>(8-44) | 28.9±3.8<br>(2-51) | $p=0.110^c$ |
| MMSE score | 29.1±0.2<br>(27-30) | 29.1±0.3<br>(26-30) | 25.9±0.8<br>(19-30) | 26.7±0.9<br>(16-30) | $p<0.001^d$ |
| MPRAGE smoothness FWHM | 2086.5±21.0<br>(1856.5-2389.5) | 2069.4±36.7<br>(1788.7-2392.6) | 2163.9±34.0<br>(1855.7-2458.8) | 2117.1±38.6<br>(1887.9-2450.1) | $p=0.289^e$ |
| Absolute head displacement (DWI) | 1.62±0.09<br>(1.30-2.77) | 1.85±0.17<br>(1.25-3.82) | 1.71±0.11<br>(1.27-3.38) | 1.61±0.08<br>(1.25-2.43) | $p=0.447^f$ |
| Relative head displacement (DWI) | 0.51±0.02<br>(0.26-0.75) | 0.48±0.03<br>(0.13-0.71) | 0.42±0.02<br>(0.28-0.59) | 0.44±0.03<br>(0.18-0.60) | $p=0.119^g$ |
| Independent validation group |  |  |  |  |  |
| sample size | 24 | 13 | 14 | 7 | -- |
| sex f/m (%) | 58.3/41.7 | 38.5/61.5 | 50.0/50.0 | 42.9/57.1 | $p=0.6545^h$ |
| age (years) | 69.7±1.4<br>(51.6-81.6) | 69.1±1.9<br>(54.1-75.8) | 70.9±1.9<br>(57.9-84.1) | 62.2±3.0<br>(53.1-75.0) | $PSP>CBS$<br>$p<0.05^i$ |
| MPRAGE smoothness FWHM | 1989.6±24.7<br>(1781.9-2238.2) | 2048.9±46.2<br>(1824.9-2380.2) | 2092.1±28.5<br>(1937.1-2266.6) | 2061.2±78.9<br>(1863.2-2364.5) | $Controls<PSP$<br>$p<0.05^j$ |
| Absolute head displacement (DWI) | 1.46±0.04<br>(1.19-1.98) | 1.47±0.08<br>(1.27-2.44) | 1.59±0.06<br>(1.31-2.01) | 1.62±0.11<br>(1.25-1.96) | $p>0.05^k$ |
| Relative head displacement (DWI) | 0.48±0.02<br>(0.23-0.70) | 0.48±0.02<br>(0.25-0.59) | 0.44±0.03<br>(0.32-0.61) | 0.48±0.05<br>(0.30-0.65) | $p>0.05^l$ |

### Cross-validation results

A summary of the cross-validation classification results obtained is presented in Figure 3. The mean and maximum accuracies were calculated over the number of features used for classification (ROIs or PCA components). The accuracy results obtained for the pairwise comparisons are all above chance level (50%), however some of these are lower than previous reports, e.g. (Haller et al., 2012; Salvatore et al., 2014), in which participants were not specifically matched for demographic and clinical features and/or motion.

**Figure 3.**
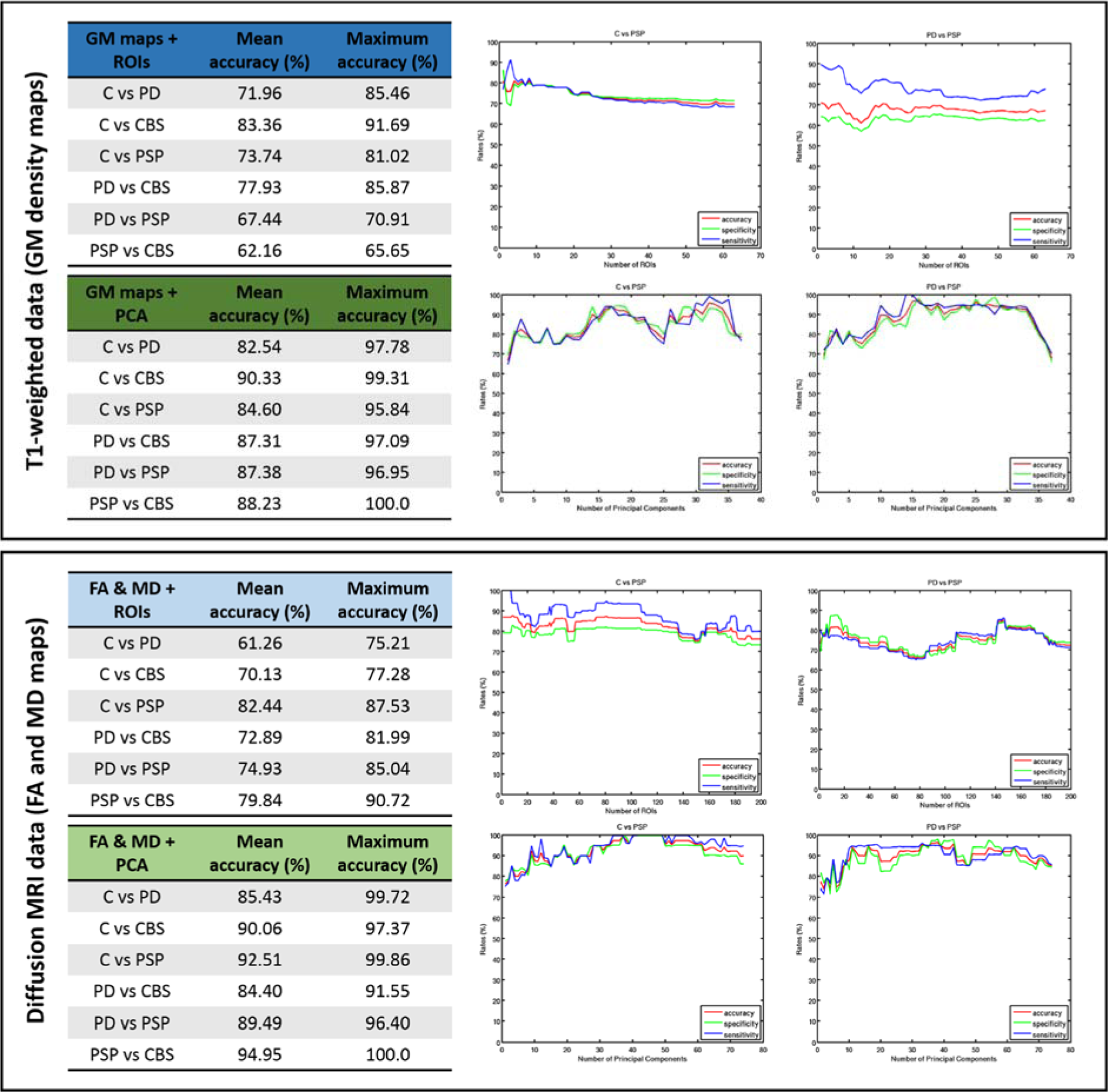
Classification accuracies achieved for pairwise comparisons using a leave-two-out cross-validation approach. For each pairwise comparison, two patients, one from each group, were left out of the training phase for each cross-validation fold and used to estimate model accuracy. The classification accuracies presented correspond to the mean and maximum accuracies obtained when different numbers of features (ROIs or PCA components) are included in the statistical model.

The strongest results were achieved when PCA was used as the method for feature extraction, resulting in mean accuracies above 80% and maximum accuracies above 90% for all pairwise comparisons, for both diffusion and T1-weighted data. The accuracies obtained with PCA were always greater than the corresponding ones obtained with ROIs for all pairwise comparisons.

For example, this difference in accuracy was 24% for the classification of controls vs PD patients using diffusion data, and 26% for the classification of PSP vs CBS patients using T1-weighted data.

The results obtained with diffusion and T1-weighted data were generally similar. However, there were also some notable differences. For example, using diffusion data and white matter ROIs, the mean classification accuracy for controls and PD patients was 61.26%, which was significantly lower that the accuracy obtained with T1-weighted data and grey matter ROIs (71.96%). In contrast, the diffusion data resulted in better differentiation between PSP and CBS patients (79.84% for diffusion data and 62.16% for T1-weighted GM maps). While neither data type outperformed the other in all cases, the T1-weighted data always resulted in higher classification accuracies for the comparisons between controls and CBS, and between PD and CBS patients. In contrast, diffusion data produced higher accuracies when comparing controls and PSP patients, and PSP and CBS patients.

Figure 3 also shows plots for classification accuracy, sensitivity and specificity as a function of the number of features included in the model (number of ROIs or PCA principal components). Sample plots for the comparisons between controls and PSP and between PD and PSP are shown as representative examples. When PCA is used for feature selection, accuracy, sensitivity and specificity generally increase as more features are added to the model until a plateau is reached at around 15 components for T1-weighted data and 15-30 for diffusion data; this level of accuracy is generally sustained until the last 2-5 features are added, which results in a decrease in classification accuracy, specificity and sensitivity. This observation is consistent with previous studies (Salvatore et al., 2014) and was to be expected since features were first selected for their ability to explain variability in the data (PCA), followed by ranking in terms of each feature’s ability to discriminate between the two subjects classes (FDR). These two levels of feature ranking ensure that noisy information is concentrated in a small number of features.

This pattern, however, is no longer observed when ROIs are used as features. This is expected, since only the FDR criterion has been used for feature ranking. FDR ranking is repeated for each cross-validation fold, and therefore the ranking of individual features will be different for each round of the cross-validation process. The ROIs were selected using anatomical criteria and therefore stay the same for each cross-validation round, while PCA features are selected for their ability to explain variance in a data-driven approach which is also repeated per cross-validation fold. For this reason the noisy information is contained in a small number of features when PCA is used, while for ROIs the noise is more evenly distributed across features.

### Independent validation results

Figure 4 shows a summary of the results obtained when the independent validation sample was used to estimate model accuracy. The mean and maximum accuracies were calculated over the number of features used for classification (ROIs or PCA components).

**Figure 4.**
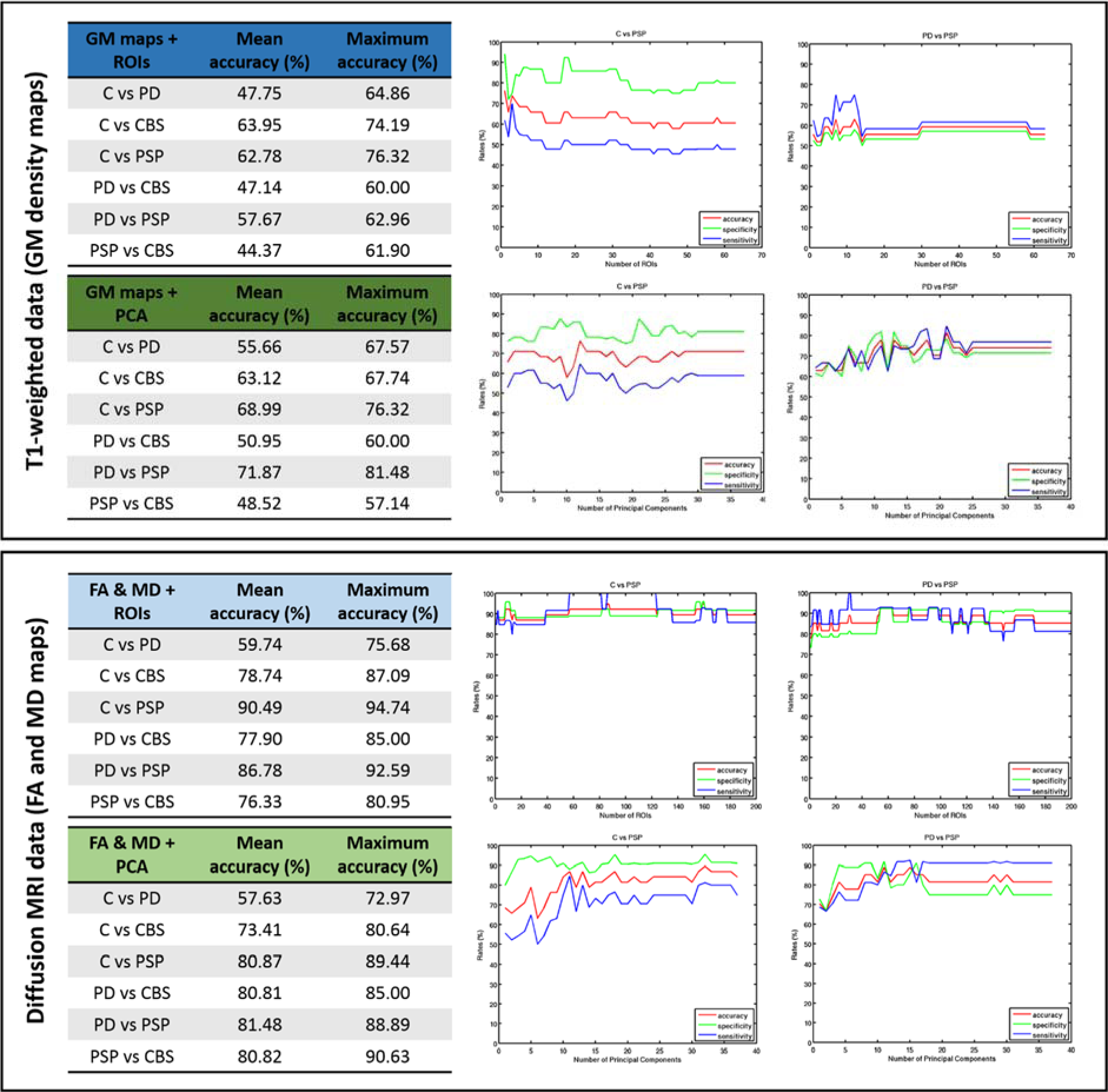
Classification accuracies achieved using the independent validation group. 76 subjects (19 from each group) were used to train the model, and validation was performed on 58 unseen patients and controls. The classification accuracies presented correspond to the mean and maximum accuracies obtained when different numbers of features are included in the statistical model (ROIs or PCA components).

Training and testing in less well matched independent sets of subjects resulted in mean classification accuracies in the range 44.37-71.87% for T1-weighted data and 57.63-90.49% for diffusion data. This decrease in classification accuracy partly reflects the overestimation inherent to a cross-validation approach, but may also reflect less stringent matching (Table 1). However, when diffusion metrics (FA and MD) were used the decrease in accuracy was less marked, and in some cases the accuracy was actually higher in the independent dataset when ROIs were used as features. The average decrease in mean classification accuracy from cross-validation to independent samples for diffusion data was 10.9% (range: 1.52 to 27.08%). For the comparisons between controls and CBS, controls and PSP, PD and CBS, and PD and PSP the mean classification accuracy in the independent sample increased by 8.61%, 8.05%, 5.01% and 11.85%, respectively, when compared to the cross-validation results.

For T1-weighted GM maps, the mean classification accuracies in the independent validation group decreased on average 22.85% (range: 9.77 to 39.71%), and the accuracies achieved were no longer significantly above chance for several group comparisons. The decrease in mean classification accuracy was more accentuated when PCA was used for feature selection.

Figure 4 also shows sample representative plots for classification accuracy, sensitivity and specificity as a function of the number of features included in the model (number of ROIs or

PCA principal components). In some cases there was an initial increase in classification accuracy, sensitivity and specificity as more features were added to the model, but in general the results obtained were very stable and independent from the number of features used.

## Discussion

Our study addresses four key issues in the use of MRI for diagnostic or classification biomarkers for parkinsonian disorders. We show that even with well-matched groups of equal size, and with control of differential motion artefacts, machine leaning with cross-validation provides accurate differential diagnosis of PD, PSP and CBS. Good diagnostic accuracy can be achieved using either grey or white matter features from standard structural and diffusion MRI sequences respectively, but diffusion-weighted images provided better generalisaiton to an independent validation dataset. Using a principal components analysis over grey or white matter provides higher classification accuracies compared to a set of anatomical regions-of-interest.

Close matching by demographics, clinical severity and motion artefacts is essential to properly evaluate and compare candidate biomarkers. Without such matching, the apparent success of some previous imaging-based biomarkers in distinguishing clinical groups may have been inflated by individual differences that are unrelated to the structural and neuropathological consequences of disease. For example, in unselected cases, motion artefacts were greater in patients than controls: 26% of patients exceeded our motion criteria compared to only 10% of controls. Differences were also observed between patient groups: 9% of PD patients (3 of 35) were excluded, compared to 28% of CBS patients (10 of 36) and 37% of PSP (19 of 52) patients.

Machine learning tools such as support vector machines are very sensitive to systematic patterns in the data but are agnostic as to the origins of such patterns e.g. motion *versus* neuropathology *versus* atrophy. The very high classification accuracies between patient groups reported in previous studies (100% in some studies), may have been inflated by different levels of motion. The effects of head motion in MRI data analysis are well documented in the literature. For example, head motion during acquisition of 3D T1-weighted MRI images results in reduced grey matter volume estimates (Reuter et al., 2015), while head motion in a diffusion MRI acquisition can result in spurious group differences in diffusion metrics (Yendiki, Koldewyn, Kakunoori, Kanwisher, & Fischl, 2014). Similarly, the comparison of groups at different stages of disease, or different levels of severity, would confound group-membership with severity. Unfortunately, there is no universal severity or staging rating scale across parkinsonian disorders. The PSP-rating-scale (Golbe & Ohman-Strickland, 2007), and a new CBS-rating-scale under development include disease specific clinical features, but we applied the UPDRS-III with its focus on common motor features across our three clinical groups.

The selection of features is critical to the performance and interpretation of classifiers. MRI provides a rich repertoire of structural, functional, neurochemical and diffusion features. We focus on the T1-image and diffusion tensor images which are most widely available, with short sequences that are readily tolerated by patients, and which require minimal operator expertise. These would be an advantage for scalable multisite studies, or in support of diagnostics and stratification in a trial context. Nonetheless, even these standard sequences provide many potential features and feature extraction options.

We compared two approaches for feature extraction, based on (i) *a priori* regions of interest from a common anatomical atlas, and (ii) a data driven approach using a principal components analysis across subjects. Our cross-validation results suggest that using principal components analysis over the full extent of grey or white matter voxels provides higher classification accuracies when compared to calculating mean values over a set of anatomical ROIs (mean accuracies were on average 14 percentage points higher for PCA features than ROIs with T1-weighted data and 16 points for diffusion data). This advantage of PCA could be due to small localised changes in brain morphology and/or function that are averaged across a ROI. On the other hand, the differences between the two feature extraction methods are significantly reduced when an independent sample is used for validation. This suggests that the PCA approach may be more vulnerable to the overfitting with cross-validation approaches.

We also compared two types of feature – GM density measures based on a T1-weighted sequence and metrics of white matter tissue organisation using diffusion tensor imaging. Our results replicated previous studies in showing that both types of data result in classification accuracies significantly above the chance level. Neither feature type clearly outperforms the other across all pairwise comparisons among our three clinical cross-validation groups.

However, features obtained from diffusion MRI data resulted in significantly higher classification accuracies when an independent validation cohort, for both methods of feature extraction (ROIs and PCA). For some contrasts (controls vs CBS, controls vs PSP, PD vs CBS, and PD vs PSP) the classification accuracy in the independent sample using diffusion data was as good as the cross-validation results.

The issue of disease severity is challenging, from two perspectives. First, there is currently no single rating scale or investigation that fully summarises disease severity across PD, PSP and CBS, either as a clinical scale, neurotransmitter or functional brain image. Even where a clinical scale such as UPDRS is applicable across the disorders, it may not give a like for like comparison in terms of disease stage (e.g., from onset to death) or functional decline (e.g., activities of daily living), or pathology (e.g., dopamine depletion, or cell loss). Second, the three diseases may each have prolonged prodromal phases and long periods in which patients are misdiagnosed. PSP and CBD typically take 2.5-3 years from symptoms to diagnosis (Coyle-Gilchrist et al., 2016; Mamarabadi, Razjouyan, & Golbe, 2018), while PD causes under-recognised clinical manifestations like constipation and REM-sleep behavioural disorder many years before tremor and akinesia. It is too soon to know whether MRI based classification is capable of differentiating these disorders in the early prodromal stages, or even pre-symptomatically, in the way that has been shown for frontotemporal dementia (Rohrer et al., 2015). The recent operationalization of early stage ‘oligosymptomatic’ cases, and ‘possible’ versus ‘probable’ cases will enable MRI biomarkers of PSP to be tested earlier (MDS criteria (Höglinger et al., 2017)).

Phenotypic variation other than severity is also a challenge. The classical presentation of PSP, as Richardson’s syndrome, has very high clinico-pathological correlations to PSP-pathology. However, in recent years it has been shown that this classical phenotype represents a minority of cases of PSP-pathology: cognitive, linguistic and behavioural presentations are common (Höglinger et al., 2017; Respondek et al., 2014). Similarly, CBS has many phenotypic variants, with motoric, behavioural and language presentations (Armstrong et al., 2013). This study does not include cases from the full phenotypic range of corticobasal syndromes, or syndromes caused by corticobasal degeneration (Alexander et al., 2014). The current study was not designed to resolve the issue of heterogeneity, but rather to highlight methodological considerations, and best practice, which we hope can be carried forward to identify robust biomarkers of a wide range of phenotypic expressions of the pathologies of PD, PSP and corticobasal degeneration.

Although we attempted to address the methodological limitations of previous studies, some limitations remain. This was a single centre study, resulting in a modest sample size when compared to recent multi-centre studies (Huppertz et al., 2016; Nigro et al., 2017). This limits the generalisation of our results to different clinical sites with potentially different scanning practices, scanner manufacturers and sequence parameters. The control and patient groups included in this study were matched for age, sex, motion parameters and UPDRS-III scores (for the patients). However, there was a difference in MMSE between the groups; and previous studies have highlighted the cognitive impairments resulting from PD (Williams-Gray et al., 2013), PSP (Rittman et al., 2013), and CBS (Burrell, Hodges, & Rowe, 2014). Given the correlations between cognitive function and structural and diffusion MRI in Parkinson’s disease, PSP-Richardson’s syndrome, and CBS (Ghosh et al., 2012; Mak et al., 2015; Paviour et al., 2006; Rae et al., 2012) the non-matching by cognitive dysfunction could contribute to classification. Against this argument, is that different cognitive deficits are hallmarks of PD, PSP and CBS, and to match a cognitive profile would compromise the representativeness of the patients chosen.

Another potential limitation of this study is that there is not always correspondence between current clinical diagnosis and neuropathology at post-mortem. Our patient labels were assigned using clinical diagnostic criteria not histopathology, and therefore might not be perfectly defined and independent, capping the statistical classifier’s ability to learn and separate the different patterns of disease. Our centre’s diagnostic accuracy of CBS and PSP is in line with other centres (Alexander et al., 2014; Gazzina et al., 2019), with generally high clinicopathological correlation of PSP-Richardson’s syndrome (>90%) relative to CBS/CBD (>60%). Finally, all our data were subjected to strict data quality control criteria, with the aim that the disease patterns detected by SVM were independent of the severity of motion present in those data. While this ensures that poor data quality will not be mistaken for real effects of the pathology, it may also exclude patients with symptoms that do not allow them to be still enough to undergo the MRI examination. For example, 19 subjects with PSP (37% of the original sample) were excluded by our quality control criteria, which may bias the sample in the PSP group. This means the patient sample included in our analysis may not be representative of the full range of disease.

In summary, we suggest that machine learning methods for MRI data can be used to aid the automatic differential diagnosis of PSP, CBS and PD, meeting critical critieria set by the Movement Disorder Society Neuroimaging Study Group and the JPND Working group ASAP-SYn-Tau (van Eimeren et al., 2019). However to make such a contribution, and augment clinical assessments, these techniques must guard against methodological biases from different levels of motion across patient groups, and poorly matched samples. With closely matched groups, of equal size and similar severity, the use of diffusion weighted images is particularly encouraging, in its high accuracy rate and generalization to independent data. Application of these methods to large samples and multisite studies will be facilitated by international collaborative studies of early stage or atypical presentations of each disease (eg PROSPECT-UK (Woodside et al., 2017) and the Four-repeat tauopathy neuroimaging initiative), aiming for reliable, unbiased, disseminated tools for early differential diagnosis and stratification in clinical trials of new therapies.

## Data Availability

Participant consent prevents open data access but academic (non-commercial) requests for data sharing would be welcome. Please contact the senior author.

